# Experience of clinical services shapes attitudes to mental health data sharing: Findings from a UK-wide survey

**DOI:** 10.1101/2021.02.02.21251041

**Authors:** E.J. Kirkham, S. M. Lawrie, C.J. Crompton, M.H. Iveson, N. D. Jenkins, J. Goerdten, I. Beange, S.W.Y. Chan, A. McIntosh, S. Fletcher-Watson

**Affiliations:** Division of Psychiatry, Centre for Clinical Brain Sciences, University of Edinburgh, Kennedy Tower, Royal Edinburgh Hospital, Morningside Park, Edinburgh, EH10 5HF, UK; Edinburgh Dementia Prevention & Centre for Clinical Brain Sciences, University of Edinburgh, Edinburgh, UK; Department of Epidemiological Methods and Etiological Research, Leibniz Institute for Prevention Research and Epidemiology–BIPS, Bremen, Germany; Department of Clinical Psychology, School of Health in Social Science, University of Edinburgh, UK

**Author notes:** **Corresponding author details:** Division of Psychiatry, Centre for Clinical Brain Sciences, University of Edinburgh, Kennedy Tower, Royal Edinburgh Hospital, Morningside Park, Edinburgh, EH10 5HF, UK.

**Keywords:** mental health, data sharing, health data, patient satisfaction, patient perspectives

## Abstract

**Background:** Routinely-collected mental health data could deliver novel insights for mental health research. However, patients’ willingness to share their mental health data remains largely unknown. We investigated factors influencing likelihood of sharing these data for research purposes amongst people with experience of mental illness.

**Methods:** We collected responses from a diverse sample of UK National Health Service (NHS) users (n = 2187) of which about half (n = 1087) had lifetime experience of mental illness. Ordinal logistic regression was used to examine the influence of demographic factors, clinical service experience, and primary mental illness on willingness to share mental health data, contrasted against physical health data.

**Results:** There was a high level of willingness to share mental (89.7%) and physical (92.8%) health data for research purposes. Higher levels of satisfaction with the NHS were associated with greater willingness to share mental health data. Furthermore, people with personal experience of mental illness were more willing than those without to share mental health data, once the effect of NHS satisfaction had been controlled for. Of the mental illnesses recorded, people with depression, obsessive-compulsive disorder (OCD), personality disorder or bipolar disorder were significantly more likely to share their mental health data than people without mental illness.

**Conclusions:** These findings suggest that positive experiences of health services and personal experience of mental illness are associated with greater willingness to share mental health data. NHS satisfaction is a potentially modifiable factor that could foster public support for increased use of NHS mental health data in research.

## Background

Routinely-collected health data, such as those held by the UK’s National Health Service (NHS) in electronic health records, are highly valuable for mental health research. Not only do these data represent millions of individuals, spanning entire life courses, they also capture information from people who are simultaneously more in need of improved treatments and less likely to participate in traditional research studies (1).

However, attempts to increase the sharing of such routinely collected data can suffer catastrophic failure if they are not supported by the public who provide the data, as demonstrated by the scandal surrounding NHS England’s attempt to share health data on an opt-out basis in the early-2010s (2). Whilst projects using routinely-collected health data do engage with special panels designed to approximate public opinion (e.g. the Public Benefit and Privacy Panel in Scotland; (3)), and there is a growing literature examining the general public’s views on sharing routinely-collected health data (4), there have been no large-scale quantitative investigations of the views of people with personal experience of mental illness. Indeed, whilst there is evidence that the general public as a whole views mental health data as more sensitive than other forms of health data (4-8), it remains unclear whether this view is shared by people with lived experience of mental illness. Not only does this result in stakeholders’ priorities remaining potentially unmet, it also means that researchers’ particular difficulties in accessing mental health data (9) could be partly based on unsubstantiated assumptions.

The limited data available tend to suggest that the organisation that will process the mental health data and the perceived sensitivity of these data are important factors in people’s decisions about data sharing (10-13). However, without larger, more representative samples it remains impossible to ascertain what characteristics and experiences affect willingness to share mental health data, and how this contrasts with willingness to share physical health data. This contrast is important given that most studies of this nature focus primarily on physical health data, and it remains unknown whether their findings translate to mental health data, which may be more stigmatised (14).

To this end, we conducted a UK-wide online survey of adult NHS-users, to examine which factors influence willingness to share routinely-collected mental health data, with a focus on the views of people with experience of mental illness. We recruited a diverse sample to examine the influence of demographic factors, and embedded the work within the context of the NHS, the primary source of routinely-collected health data in the UK.

## Methods

### Study design and population

A first draft of the survey was designed by the research team. This draft was presented to a group of people with lived experience of mental illness, and changes to wording and layout were made based on their feedback. The survey was also sent to research clinicians and further suggested alterations were incorporated. The survey covered participants’ views about sharing mental and physical health data, their personal experience of mental and physical health, and their demographic information. The median response time was 11 minutes. The final survey was administered online through the Qualtrics platform (15) and the full script is available at https://doi.org/10.7488/ds/3146 and in Appendix 1. The research received ethical approval from the School of Health in Social Sciences Research Ethics Committee at the University of Edinburgh.

Participants were recruited from December 2018 to August 2019 via social media, posters, and in-person at a science festival. A number of research and clinical teams around the UK promoted the survey through their networks, but there was no direct recruitment of patients via clinical settings. This strategy formed the basis of our recruitment due to financial and time constraints. However, we also sought to extend the reach of the survey where possible, by working with a company to advertise the survey on Facebook. We ensured that a portion of this advertising was presented specifically to men and to people without university degrees, as these demographic groups were under-represented following our initial recruitment. In addition, we invited people from less represented groups to take part using the study recruitment website Prolific Academic (16).

All participants took part voluntarily; 174 individuals who took part through Prolific Academic were paid approximately £2. These participants were primarily from Black, Asian or Minority Ethnic groups: we targeted these populations to improve the ethnic diversity of the sample. All participants were over 16 and stated that they had used the NHS for either physical or mental health care. Participants were classified as having experience of mental illness if they self-reported that they had had a mental illness at some time in their life (diagnosis was not required). Participant demographics are presented in Table 1. A total of 2187 participants contributed data to the study. However, the number of participants responding to each question varied, due to participant drop-out and built-in exclusion criteria (e.g. people were not asked specific questions about treatment for mental illness if they reported no experience of mental illness). Consequently, the number of participants differs between analyses.

**Table 1.** Participant demographics

|  | All participants | Experience of mental illness (ever) |  |  |
| --- | --- | --- | --- | --- |
|  |  | Yes | No | Prefer not to say |
| Total, n | 2187 | 1087 (65.09%) | 529 (31.68%) | 54 (3.23%) |
| Gender, n (%) |  |  |  |  |
| Male | 514 (31.28%) | 314 (29.51%) | 187 (35.82%) | 12 (22.64%) |
| Female | 1094 (66.59%) | 721 (67.76%) | 334 (63.98%) | 36 (67.92%) |
| Non-binary or prefer to self-describe | 19 (1.16%) | 18 (1.69%) | 0 (0.00%) | 1 (1.89%) |
| Prefer not to say | 16 (0.97%) | 11 (1.03%) | 1 (0.19%) | 4 (7.55%) |
| Location, n (%) |  |  |  |  |
| England | 1050 (63.99%) | 695 (65.38%) | 316 (60.77%) | 36 (67.92%) |
| Scotland | 490 (29.86%) | 300 (28.22%) | 174 (33.46%) | 14 (26.42%) |
| Wales | 65 (3.96%) | 49 (4.61%) | 15 (2.88%) | 1 (1.89%) |
| Northern Ireland | 19 (1.16%) | 10 (0.94%) | 9 (1.73%) | 0 (0.00%) |
| Outside UK | 17 (1.04%) | 9 (0.85%) | 6 (1.15%) | 2 (3.77%) |
| Ethnicity, n (%) |  |  |  |  |
| White | 1438 (88.38%) | 961 (91.26%) | 431 (83.37%) | 41 (80.39%) |
| Asian/Asian British | 92 (5.65%) | 38 (3.61%) | 51 (9.86%) | 3 (5.88%) |
| Black/African/Caribbean/Black British | 53 (3.26%) | 24 (2.28%) | 26 (5.03%) | 2 (3.92%) |
| Mixed/multiple ethnic groups | 32 (1.97%) | 21 (1.99%) | 7 (1.35%) | 4 (7.84%) |
| Other | 12 (0.74%) | 9 (0.85%) | 2 (0.39%) | 1 (1.96%) |
| Highest education, n (%) |  |  |  |  |
| Postgraduate degree or professional qualification | 587 (36.01%) | 383 (36.23%) | 181 (34.94%) | 21 (42.00%) |
| Undergraduate degree | 390 (23.93%) | 260 (24.60%) | 122 (23.55%) | 8 (16.00%) |
| Vocational or college qualification | 246 (15.09%) | 147 (13.91%) | 91 (17.57%) | 6 (12.00%) |
| A-levels or equivalent | 189 (11.60%) | 121 (11.45%) | 62 (11.97%) | 5 (10.00%) |
| GCSEs or equivalent | 200 (12.27%) | 135 (12.77%) | 55 (10.62%) | 10 (20.00%) |
| Primary school | 18 (1.10%) | 11 (1.04%) | 7 (1.35%) | 0 (0.00%) |
| Experience of physical disability or long-term physical illness (ever), n (%) |  |  |  |  |
| Yes | 871 (52.56%) | 598 (55.84%) | 244 (46.21%) | 27 (50.94%) |
| No | 774 (46.71%) | 468 (43.70%) | 283 (53.60%) | 20 (37.74%) |
| Prefer not to say | 12 (0.72%) | 5 (0.47%) | 1 (0.19%) | 6 (11.32%) |
| Age (years), mean (SD) | 45.58 (17.38) | 44.44 (17.03) | 48.26 (17.74) | 42.14 (18.18) |

Willingness to share both mental and physical health data for research was first captured using a binary (yes/no) scale. Second, to analyse predictors of data sharing, the two primary outcome measures were taken from the question “How likely would you be to share mental/physical health data for research purposesã” Each response was measured on a Likert scale from 1 (extremely unlikely) to 5 (extremely likely). The survey assessed satisfaction with the NHS using four items, covering satisfaction with first contact for a mental/long-term physical health condition and satisfaction with mental/physical health care during the previous 12 months. Each item was measured on a 5-point Likert scale from “very dissatisfied” to “very satisfied” (Appendix 1). For analysis, one NHS satisfaction variable was created from a mean of these four responses (17). Cronbach’s alpha for this new variable was 0.71 (18), indicating acceptable internal consistency.

### Statistical analysis

All analyses were performed in IBM SPSS Statistics for Windows, Version 24. In cases where ordinal logistic regression with proportional odds was used, the assumption of proportional odds was tested using a full likelihood ratio test.

#### Likelihood of sharing health data

McNemar’s and Wilcoxon signed-ranks tests were used to examine whether there were significant differences in willingness to share mental and physical health data amongst the full sample and amongst those with experience of mental illness. Following this, a Mann-Whitney U test was used to examine whether likelihood of sharing mental or physical health data differed between people with experience of mental illness only and people with experience of long-term physical illness only.

#### Factors influencing likelihood of sharing health data

##### Influence of demographic factors (Model 1, Table 2)

Cumulative odds ordinal logistic regression with proportional odds was used to determine the effect of demographic factors on likelihood of sharing health data (Model 1). The factors included in Model 1 were based on theoretical consideration of the topic, and were as follows: gender, age, ethnicity, location, education, experience of mental illness (ever), experience of long-term physical illness or disability (ever), self-rating of current mental health and self-rating of current physical health. The aforementioned factors were used to predict two dependent variables: (a) likelihood of sharing mental health data and (b) likelihood of sharing physical health data (Models 1a and 1b, Table 2). Age was treated as a continuous variable, self-rating of current mental/physical health was treated as ordinal (measured on a scale from 1 to 5), and the remaining variables were treated as categorical (reference categories are included in the Notes section of Table 2). Response categories for these variables are detailed in Table 1, and details of the questions can be found in Appendix 1. Being paid for survey participation was included as a co-variate. Likelihood of sharing mental/physical health data was measured on an ordinal scale from 1 to 5.

**Table 2.**
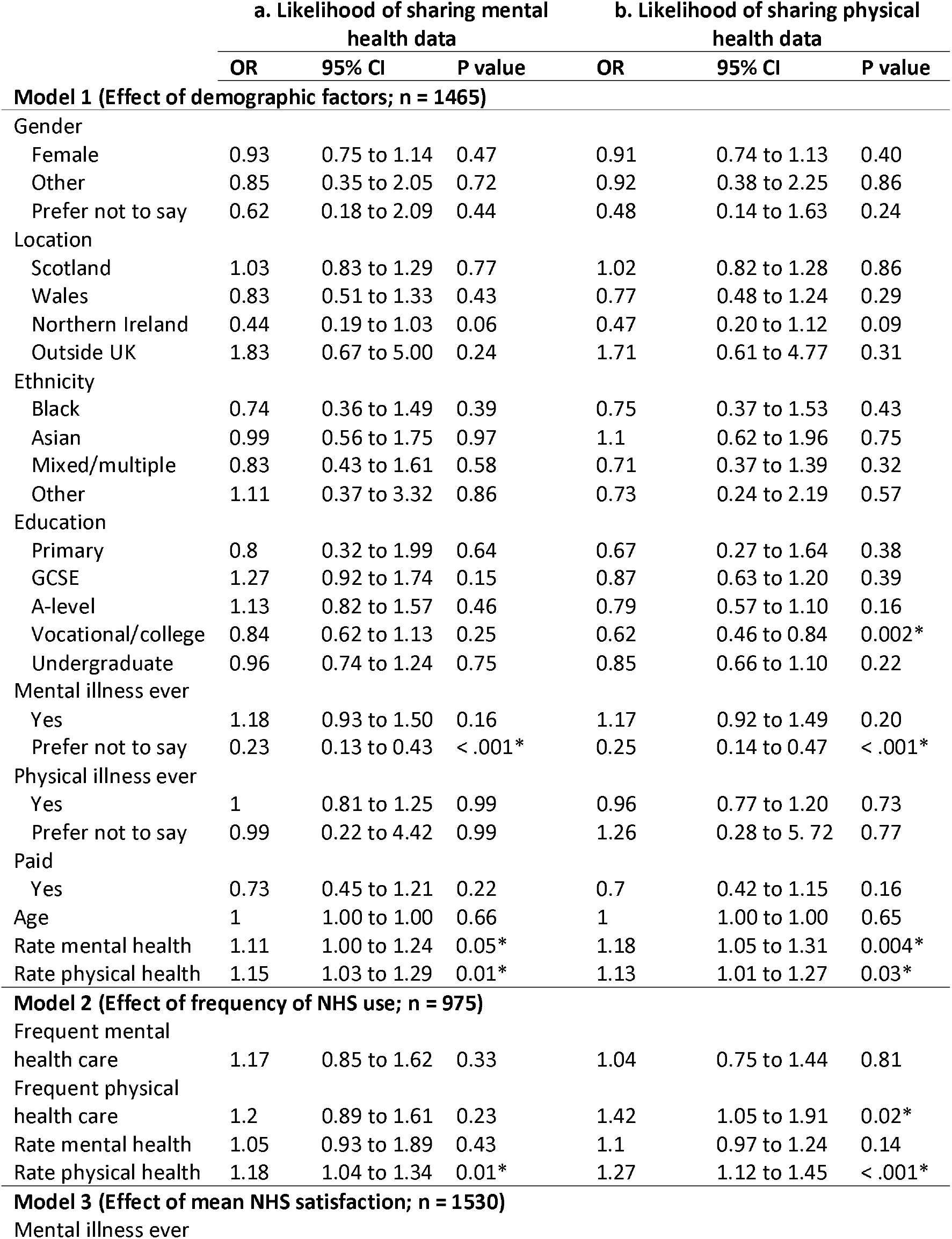
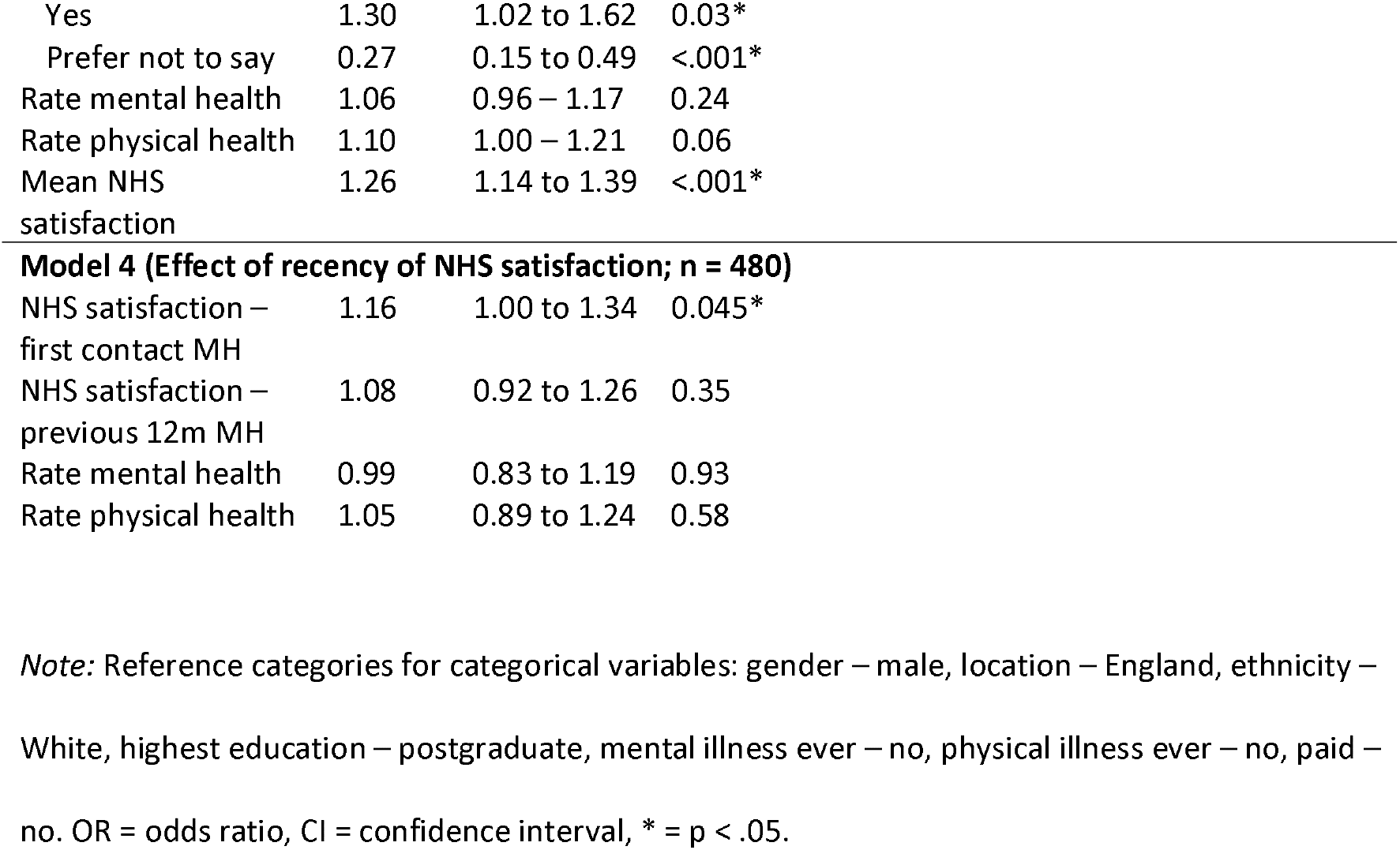
Ordinal logistic regression models examining factors affecting likelihood of sharing (a) mental and (b) physical health data

|  | a. Likelihood of sharing mental health data |  |  | b. Likelihood of sharing physical health data |  |  |
| --- | --- | --- | --- | --- | --- | --- |
|  | OR | 95% CI | P value | OR | 95% CI | P value |
| **Model 1 (Effect of demographic factors; n = 1465)** |  |  |  |  |  |  |
| Gender |  |  |  |  |  |  |
| Female | 0.93 | 0.75 to 1.14 | 0.47 | 0.91 | 0.74 to 1.13 | 0.40 |
| Other | 0.85 | 0.35 to 2.05 | 0.72 | 0.92 | 0.38 to 2.25 | 0.86 |
| Prefer not to say | 0.62 | 0.18 to 2.09 | 0.44 | 0.48 | 0.14 to 1.63 | 0.24 |
| Location |  |  |  |  |  |  |
| Scotland | 1.03 | 0.83 to 1.29 | 0.77 | 1.02 | 0.82 to 1.28 | 0.86 |
| Wales | 0.83 | 0.51 to 1.33 | 0.43 | 0.77 | 0.48 to 1.24 | 0.29 |
| Northern Ireland | 0.44 | 0.19 to 1.03 | 0.06 | 0.47 | 0.20 to 1.12 | 0.09 |
| Outside UK | 1.83 | 0.67 to 5.00 | 0.24 | 1.71 | 0.61 to 4.77 | 0.31 |
| Ethnicity |  |  |  |  |  |  |
| Black | 0.74 | 0.36 to 1.49 | 0.39 | 0.75 | 0.37 to 1.53 | 0.43 |
| Asian | 0.99 | 0.56 to 1.75 | 0.97 | 1.1 | 0.62 to 1.96 | 0.75 |
| Mixed/multiple | 0.83 | 0.43 to 1.61 | 0.58 | 0.71 | 0.37 to 1.39 | 0.32 |
| Other | 1.11 | 0.37 to 3.32 | 0.86 | 0.73 | 0.24 to 2.19 | 0.57 |
| Education |  |  |  |  |  |  |
| Primary | 0.8 | 0.32 to 1.99 | 0.64 | 0.67 | 0.27 to 1.64 | 0.38 |
| GCSE | 1.27 | 0.92 to 1.74 | 0.15 | 0.87 | 0.63 to 1.20 | 0.39 |
| A-level | 1.13 | 0.82 to 1.57 | 0.46 | 0.79 | 0.57 to 1.10 | 0.16 |
| Vocational/college | 0.84 | 0.62 to 1.13 | 0.25 | 0.62 | 0.46 to 0.84 | 0.002* |
| Undergraduate | 0.96 | 0.74 to 1.24 | 0.75 | 0.85 | 0.66 to 1.10 | 0.22 |
| Mental illness ever |  |  |  |  |  |  |
| Yes | 1.18 | 0.93 to 1.50 | 0.16 | 1.17 | 0.92 to 1.49 | 0.20 |
| Prefer not to say | 0.23 | 0.13 to 0.43 | < .001* | 0.25 | 0.14 to 0.47 | < .001* |
| Physical illness ever |  |  |  |  |  |  |
| Yes | 1 | 0.81 to 1.25 | 0.99 | 0.96 | 0.77 to 1.20 | 0.73 |
| Prefer not to say | 0.99 | 0.22 to 4.42 | 0.99 | 1.26 | 0.28 to 5.72 | 0.77 |
| Paid |  |  |  |  |  |  |
| Yes | 0.73 | 0.45 to 1.21 | 0.22 | 0.7 | 0.42 to 1.15 | 0.16 |
| Age | 1 | 1.00 to 1.00 | 0.66 | 1 | 1.00 to 1.00 | 0.65 |
| Rate mental health | 1.11 | 1.00 to 1.24 | 0.05* | 1.18 | 1.05 to 1.31 | 0.004* |
| Rate physical health | 1.15 | 1.03 to 1.29 | 0.01* | 1.13 | 1.01 to 1.27 | 0.03* |
| **Model 2 (Effect of frequency of NHS use; n = 975)** |  |  |  |  |  |  |
| Frequent mental health care | 1.17 | 0.85 to 1.62 | 0.33 | 1.04 | 0.75 to 1.44 | 0.81 |
| Frequent physical health care | 1.2 | 0.89 to 1.61 | 0.23 | 1.42 | 1.05 to 1.91 | 0.02* |
| Rate mental health | 1.05 | 0.93 to 1.89 | 0.43 | 1.1 | 0.97 to 1.24 | 0.14 |
| Rate physical health | 1.18 | 1.04 to 1.34 | 0.01* | 1.27 | 1.12 to 1.45 | < .001* |
| **Model 3 (Effect of mean NHS satisfaction; n = 1530)** |  |  |  |  |  |  |
| Mental illness ever |  |  |  |  |  |  |

|  |  |  |  |
| --- | --- | --- | --- |
| Yes | 1.30 | 1.02 to 1.62 | 0.03* |
| Prefer not to say | 0.27 | 0.15 to 0.49 | <.001* |
| Rate mental health | 1.06 | 0.96 – 1.17 | 0.24 |
| Rate physical health | 1.10 | 1.00 – 1.21 | 0.06 |
| Mean NHS satisfaction | 1.26 | 1.14 to 1.39 | <.001* |

**Model 4 (Effect of recency of NHS satisfaction; n = 480)**
|  |  |  |  |
| --- | --- | --- | --- |
| NHS satisfaction – first contact MH | 1.16 | 1.00 to 1.34 | 0.045* |
| NHS satisfaction – previous 12m MH | 1.08 | 0.92 to 1.26 | 0.35 |
| Rate mental health | 0.99 | 0.83 to 1.19 | 0.93 |
| Rate physical health | 1.05 | 0.89 to 1.24 | 0.58 |

*Note:* Reference categories for categorical variables: gender – male, location – England, ethnicity –
White, highest education – postgraduate, mental illness ever – no, physical illness ever – no, paid –
no. OR = odds ratio, CI = confidence interval, \* = $p < .05$ .

##### Influence of frequency of NHS use (Model 2, Table 2)

Cumulative odds ordinal logistic regression with proportional odds was next used to determine the effect of frequency of mental and physical NHS health care on likelihood of sharing health data (Model 2). The factors included in Model 2 were frequency of NHS mental health care, frequency of NHS physical health care, self-rating of mental health and self-rating of physical health (the latter two factors were included as co-variates because they reached significance in the previous analysis examining influence of demographic factors (Model 1)). The aforementioned factors were used to predict two dependent variables: (a) likelihood of sharing mental health data and (b) likelihood of sharing physical health data (Models 2a and 2b, Table 2). Frequency of mental/physical NHS health care was treated as binary and coded as either non-frequent: less than monthly, or frequent: monthly or more. Participants who had received inpatient care were removed from this analysis, as this form of care did not fit well into a “frequency” framework. Details of the frequency questions and original response options can be found in Appendix 1.

##### Influence of satisfaction with the NHS (Models 3 and 4, Table 2)

Spearman’s rank-order correlations were run to assess the relationship between satisfaction with the NHS and likelihood of sharing mental and physical health data. In addition, Mann-Whitney U tests were used to examine whether participants with and without experience of mental illness differed in their satisfaction with the NHS.

Next, cumulative odds ordinal logistic regression with proportional odds was used to examine whether the relationship between satisfaction with the NHS and likelihood of sharing mental health data remained significant after controlling for variables which were significant in the previous regression analyses (Model 3, Table 2). To this end, Model 3 contained the following factors: mean NHS satisfaction, experience of mental illness, self-rating of mental health, and self-rating of physical health (with the latter three variables acting as co-variates). The dependent variable was likelihood of sharing mental health data.

Following this, Model 4 (Table 2) was generated to examine whether the effect of NHS satisfaction at first contact was greater than the effect of NHS satisfaction at recent (12 month) contact. Model 4 therefore contained the following factors: satisfaction with first contact with NHS mental health services, satisfaction with 12 month contact with NHS mental health services, and self-ratings of mental and physical health (as covariates). The dependent variable was likelihood of sharing mental health data.

##### Influence of specific mental health conditions

A cumulative odds ordinal logistic regression with proportional odds was run to determine the effect of primary reported mental health condition on likelihood of sharing mental health data (Table 3). Participants were asked to choose their primary mental health condition from a list provided (Appendix 1; depression, anxiety, phobia, eating disorder, schizophrenia or psychosis, OCD, personality disorder, bipolar disorder, addiction, body dysmorphic disorder, self-harm or “other”). Binary coding was used to indicate whether a given condition was the participant’s primary mental health condition or not. In light of previous findings (Table 2), NHS satisfaction and self-rating of mental/physical health were included as co-variates. This analysis was considered to be exploratory given the low number of participants whose primary mental health condition was not depression or anxiety (n = 217).

**Table 3.** Effect of mental health condition on likelihood of sharing mental health data

|  | n | Likelihood of sharing mental health data |  |  |
| --- | --- | --- | --- | --- |
|  |  | OR | 95% CI | p value |
| Primary reported mental illness |  |  |  |  |
| Depression | 450 | 1.34 | 1.03 to 1.75 | 0.03* |
| Anxiety | 352 | 1.21 | 0.91 to 1.59 | 0.18 |
| Phobia | 10 | 1.04 | 0.33 to 3.26 | 0.95 |
| Eating disorder | 29 | 1.23 | 0.61 to 2.47 | 0.57 |
| Schizophrenia or psychosis | 9 | 0.74 | 0.22 to 2.47 | 0.63 |
| Obsessive-compulsive disorder | 25 | 2.20 | 1.02 to 4.73 | 0.04* |
| Personality disorder | 23 | 2.80 | 1.24 to 6.29 | 0.01* |
| Bipolar disorder | 23 | 2.55 | 1.15 to 5.68 | 0.02* |
| Addiction or substance use disorder | 26 | 1.19 | 0.57 to 2.45 | 0.64 |
| Body dysmorphic disorder | 3 | 1.04 | 0.13 to 8.11 | 0.97 |
| Self-harm | 9 | 0.94 | 0.28 to 3.14 | 0.93 |
| Other | 60 | 1.01 | 0.61 to 1.66 | 0.98 |
| Mean NHS satisfaction |  | 1.25 | 1.13 to 1.39 | < .001* |
| Rate mental health |  | 1.08 | 0.97 to 1.19 | 0.17 |
| Rate physical health |  | 1.09 | 0.98 to 1.20 | 0.10 |

*Note.* Reference category for primary reported mental illness was no mental illness (n = 456). OR =
odds ratio, CI = confidence interval, \* = p < .05.

## Results

### Likelihood of sharing health data

Across the whole sample there was a high level of willingness to share both mental (89.68%) and physical (92.75%) health data when measured on a binary (yes/no) scale. A McNemar’s test demonstrated that this difference was significant (χ^2^(1) = 47.67, p < .001). A similar pattern was seen when dividing the sample by individuals who had experienced a mental health condition and individuals who had never experienced a mental health condition (Figure 1); both participants with experience of mental illness (χ^2^(1) = 20.02, p < .001), and participants without experience of mental illness (exact McNemar’s test p = .001) were significantly more likely to share physical health data than mental health data.

**Figure 1:**
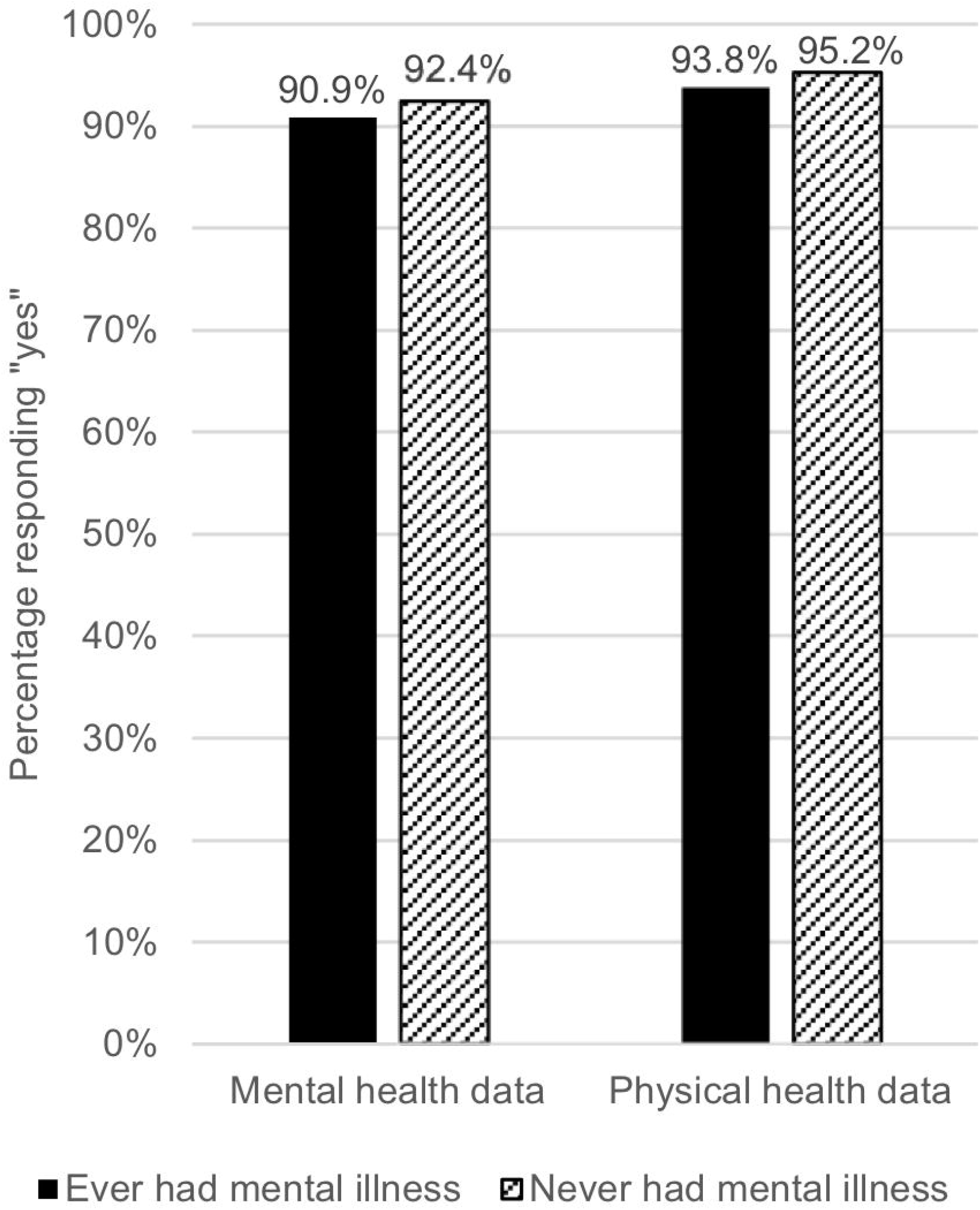
Participants who would share their mental and physical health data, split by mental health status. *Note*. n(ever had mental illness) = 1085, n(never had mental illness) = 528. Participants who responded that they would “prefer not to say” if they have ever had a mental or physical health condition are not shown.

We next examined the reported likelihood (measured on a Likert scale) of sharing mental/physical health data (Figure 2). A Wilcoxon signed-ranks test indicated that participants were significantly more likely to share their physical health data than their mental health data, z = -8.621, p < .001. This effect was also present amongst those with experience of mental illness, z = -6.412, p < .001.

**Figure 2:**
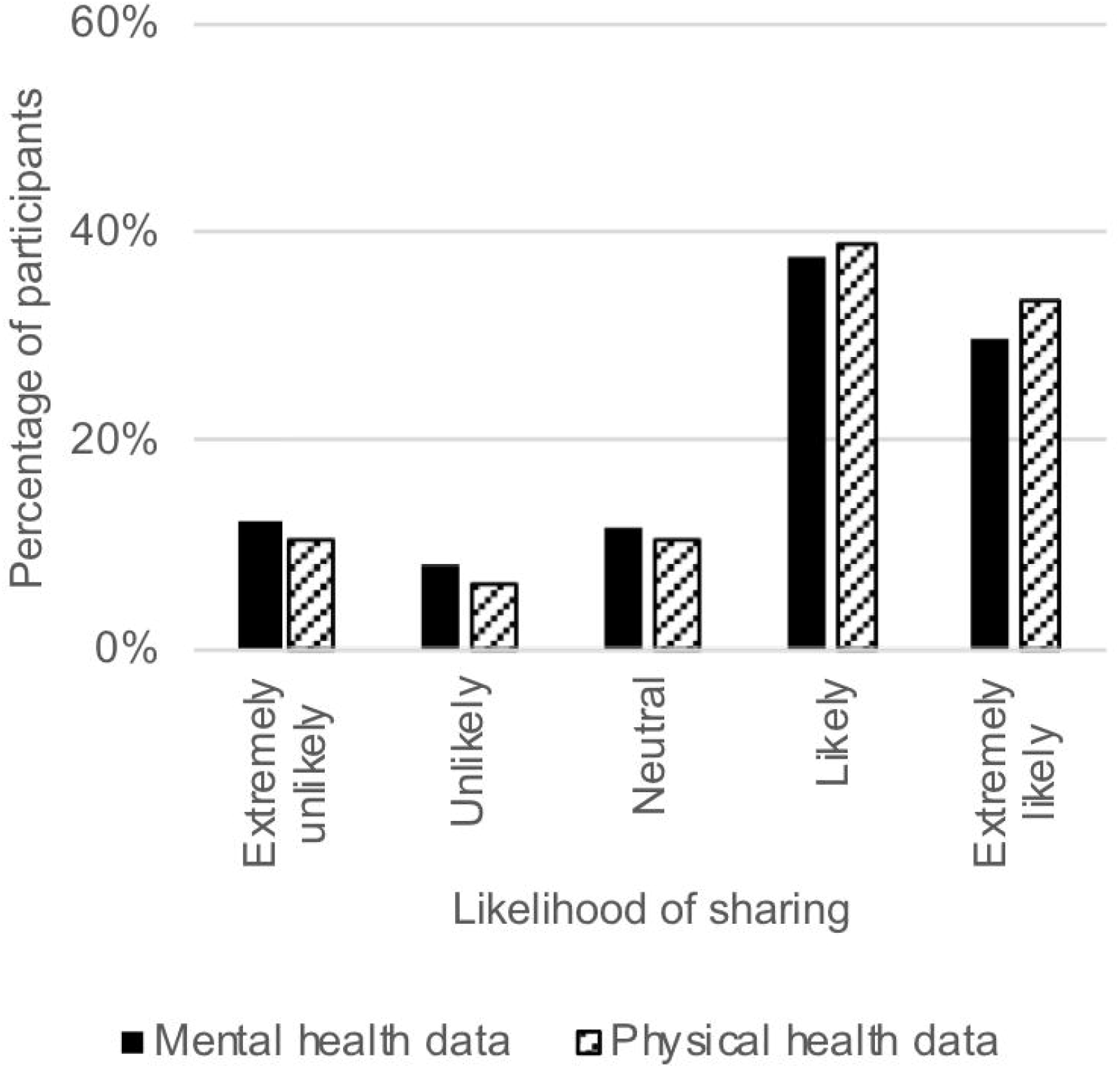
Likelihood of sharing mental and physical health data across all participants. *Note*. n = 1991

Two Mann-Whitney U tests were run to determine if likelihood of sharing mental or physical health data differed between the group of people with experience of only mental illness (n = 468) and the group of people with experience of only long-term physical illness (n = 244). Distributions of the responses for the two groups were similar, as assessed by visual inspection. There was no statistically significantly difference between people with only mental illness and people with only physical illness, for either likelihood of sharing mental health data U = 55776.50, z = -0.105, p = .917 or likelihood of sharing physical health data U = 55474.00, z = -0.327, p = .744.

### Factors influencing likelihood of sharing health data

#### Influence of demographic factors (Model 1)

The full ordinal logistic regression model examining the effect of demographic factors on likelihood of sharing mental health data (Model 1a, Table 2) significantly predicted the outcome over and above the intercept-only model χ^2^(24) = 68.80, p < .001. The assumption of proportional odds was met χ^2^(72) = 74.97, p = .38. There was no significant difference between people who had and had not experienced a mental illness in the likelihood of sharing mental health data. Those who said they would “prefer not to say” if they had ever experienced a mental illness were much less willing to share their mental health data than people who said they had never had a mental illness (OR 0.23). An increase in self-rated physical health was associated with an increase in the odds of sharing mental health data (OR 1.15). An increase in self-rated mental health was marginally associated with an increase in the odds of sharing mental health data (OR 1.11).

The full ordinal logistic regression model examining the effect of demographic factors on likelihood of sharing physical health data (Model 1b, Table 2) significantly predicted the outcome over and above the intercept-only model χ^2^(24) = 87.17, p < .001. The assumption of proportional odds was not met χ^2^(72) = 105.24, p = .006. As such, an additional adapted analysis which met the assumption was run, giving a similar outcome (Appendix 2). Although there was no overall effect of highest completed level of education on likelihood of sharing physical health data, Wald χ^2^(5) = 10.22, p = .069, the contrast between the subcategories of postgraduate degree and vocational qualification was significant, such that people with a postgraduate degree were more likely to share their physical health data than people with vocational or college-level qualifications (OR 1.61). The odds of people who said they would “prefer not to say” if they had ever experienced a *mental* illness being willing to share their *physical* health data were four times lower than that of people who said they had never had a mental illness, (OR 0.25). There was no significant difference between people who had and had not experienced a mental illness in likelihood of sharing physical health data. An increase in self-rated physical health was associated with an increase in the odds of sharing physical health data (OR 1.13). An increase in self-rated mental health was also associated with an increase in the odds of sharing physical health data (OR 1.18).

#### Influence of frequency of NHS use (Model 2)

Two cumulative odds ordinal logistic regression analyses with proportional odds were run to determine the effect of frequency of mental and physical NHS health care on willingness to share (a) mental and (b) physical health data (Models 2a and 2b, Table 2). The full model (Model 2a, Table 2) significantly predicted the likelihood of sharing mental health data over and above the intercept-only model χ^2^(4) = 10.55, p = .03. The assumption of proportional odds was met χ^2^(12) = 19.98, p = .07. An increase in self-rated physical health was associated with an increase in the odds of sharing mental health data (OR 1.18). Likelihood of sharing mental health data was not predicted by frequency of NHS mental or physical health care, or self-rated mental health.

The full model (Model 2b, Table 2) also significantly predicted likelihood of sharing physical health data over and above the intercept-only model χ^2^(4) = 22.48, p < .001. The assumption of proportional odds was met χ^2^(12) = 12.00, p = .45. People who received NHS physical health care more than monthly were more likely to share their physical health data than people who received less frequent care (OR 1.42). An increase in self-rated physical health was associated with an increase in the odds of sharing physical health data (OR 1.27). There was no association between frequency of mental health care or self-rated mental health and the likelihood of sharing physical health data.

#### Influence of satisfaction with the NHS (Models 3 and 4)

Spearman’s rank-order correlations were run to assess the relationship between satisfaction with the NHS and likelihood of sharing mental and physical health data. Higher levels of satisfaction with the NHS were associated with greater willingness to share mental health data, rs(1544) = .13, p < .001 and physical health data rs(1544) = .13, p < .001. Mann-Whitney U tests were used to examine whether participants with and without experience of mental illness differed in their satisfaction with the NHS. It was found that participants with experience of mental illness were less satisfied with their overall (mean) experience with the NHS (U = 147117.50, z = -12.47, p < .001), as well as less satisfied with their first contact for a physical health condition (U = 46305.00, -5.52, p < .001) and less satisfied with the physical health care they had received in the previous 12 months (U = 160531.00, z = -5.88, p < .001).

To disentangle the relationships between these aforementioned variables, a cumulative odds ordinal logistic regression with proportional odds (Model 3, Table 2) was run to determine the relative effects of mean NHS satisfaction and experience of mental illness on likelihood of sharing mental health data (with self-rated mental and physical health included as covariate factors). The full model significantly predicted the likelihood of sharing mental health data over and above the intercept-only model (χ^2^(5) = 66.44, p < .001). The assumption of proportional odds was met (χ^2^(15) = 21.45, p = .12). In keeping with previous analyses, satisfaction with the NHS (Wald χ^2^(1) = 20.64, p < .001), and experience of mental illness were associated with likelihood of sharing mental health data (Wald χ^2^(2) = 29.92, p < .001). Participants who would “prefer not to say” whether they had a mental illness were much less willing than people who said they had never had a mental illness to share their mental health data (OR 0.27). This means that although experience of mental illness had no effect on willingness to share mental health data when examining demographic factors only (Model 1a), parsing out the effect of NHS satisfaction (by including it as a factor in Model 3) revealed a significant relationship between experience of mental illness and higher likelihood of sharing mental health data (OR 1.30).

Following this, a cumulative odds ordinal logistic regression with proportional odds was run to determine the association between two specific measures of NHS satisfaction (satisfaction with first contact for mental health care, and satisfaction with mental health care in the previous 12 months) on likelihood of sharing mental health data amongst people who have had a mental illness (Model 4, Table 2; n = 480). Ratings of mental and physical health were included as co-variates, as they significantly predicted willingness to share mental health data in a previous analyses (Model 1a). The assumption of proportional odds was met, (χ^2^(12) = 7.604, p = .82). All variance inflation factors (VIF) values were well below 10, with the largest being 1.33, indicating that there was no multi-collinearity. The final model did not significantly predict likelihood of sharing mental health data over and above the intercept-only model, χ^2^(4) = 8.885, p = .064. However, an increase in satisfaction with first contact for mental health care was associated with an increase in the odds of sharing mental health data (OR 1.16). There was no association between satisfaction with previous 12 months’ mental health care, rating of physical health or rating of mental health and willingness to share mental health data.

#### Influence of specific mental health conditions

A cumulative odds ordinal logistic regression with proportional odds was run to determine the effect of each primary reported mental health condition on likelihood of sharing mental health data (Table 3). The full model significantly predicted the outcome over and above the intercept-only model (χ^2^(15) = 43.06, p < .001). The assumption of proportional odds was met (χ^2^(45) = 52.76, p = .20). In comparison to participants with no experience of mental illness, participants whose primary experience of mental illness was with depression, OCD, personality disorder or bipolar disorder were more willing to share their mental health data (Table 3). There were no cases where experience of a mental illness significantly reduced willingness to share mental health data.

## Discussion

This study is the first to examine factors that determine likelihood of sharing mental health data for research purposes in a large UK sample. Willingness to share mental health data was high amongst people with and without experience of mental illness, though it was nevertheless lower than willingness to share physical health data. Higher satisfaction with the NHS was strongly associated with increased likelihood of sharing mental health data. Initial analyses suggested no association between mental illness experience and likelihood of sharing mental health data, but once the contribution of NHS satisfaction was controlled for, participants with experience of mental illness were more likely to share their mental health data than those without experience of mental illness. Specifically, people whose primary mental health condition was depression, OCD, personality disorder or bipolar disorder were each significantly more likely to share their mental health data than people with no experience of mental illness.

There was a high level of willingness to share both mental (89.7% “yes”) and physical (92.8% “yes”) health data, supporting previous work which showed support for sharing health data at 73%, 81% and 90% (8, 19, 20). These findings are encouraging given the value of routinely-collected health data in health and data science research (9). At the same time, if one in ten people are unwilling to share mental health data, this could still present a significant obstacle that hinders progress towards increased availability of routine mental health data for research. Therefore researchers should not underestimate the importance of continued and sustained public engagement on the value of routine data analysis for research (21), especially since participants were less willing to share mental health data than physical health data. This reluctance signals that mental health data may be more “sensitive” than other health data (4-8), especially as it comes from participants who had already chosen to engage with mental health research. Having said this, the actual size of the difference found here (3%) suggests that mental health stigma (14) is not having a dramatic effect in this case.

A core finding of this work was that satisfaction with the NHS had a particularly notable impact on willingness to share both mental and physical health data for research purposes, echoing previous findings of a positive relationship between perceived quality of care and greater confidence in the privacy of health information (22). One speculative explanation for the present finding may be that positive experiences of health care are associated with greater trust in institutional structures which deal with health care (23), and that this trust is in turn associated with greater willingness to share health data (24). Both situations involve power differences between the individual and the institution(s), and both concern personal information. Whether participants are explicitly conflating health care and health research, or whether the association is more implicit, is a matter for future qualitative research. Either way, it is apparent that ensuring positive clinical contact and facilitating awareness of research in clinical settings could contribute to increased willingness to share health data.

Furthermore, in the present study, once the impact of NHS satisfaction was taken into account, participants with experience of mental illness were more willing to share their mental health data than those without experience of mental illness. This finding has important implications for clinicians, as it suggests that experience of mental illness is not in itself a barrier to data sharing, rather it is how those with mental illness experience the NHS that matters. In addition, individuals’ satisfaction with their first contact with the NHS for mental health care may be particularly important in predicting their likelihood of sharing mental health data. The related finding that people with experience of mental illness are less satisfied with NHS care echoes previous findings of a relationship between higher depression scores and lower satisfaction with physical health care (25, 26). Reduced satisfaction could be related to diagnostic overshadowing, in which people with mental illness are less likely to receive appropriate treatment for comorbid physical illness (27, 28). Future work should examine the relationship between mental health and satisfaction with health care in further detail.

To our knowledge, this study was the first to examine the effect of experience of different mental health conditions on willingness to share health data. People whose primary mental health condition was depression, OCD, personality disorder or bipolar disorder were significantly more likely to share their mental health data than those without experience of mental illness, when confounders were taken into account. People whose primary condition was anxiety, phobia, eating disorder, psychosis, addiction, body dysmorphic disorder or self-harm did not differ from people without experience of mental illness in their willingness to share mental health data. Notably, none of the mental health conditions measured were associated with a significant reduction in likelihood of sharing mental health data. These findings should be considered in light of the small sample sizes in most of the groups; further research is needed to examine whether the present pattern of relationships between participants’ primary mental health conditions and their willingness to share mental health data remains once larger numbers of people are included. Nevertheless, taken together, people with experience of mental illness are more willing than those without experience of mental illness to share mental health data, possibly because they are more motivated to contribute towards improved treatments for mental illness (11). Interestingly, there was very little evidence of relationships between demographic factors and willingness to share mental health data, despite our recruitment of a demographically diverse UK sample with a wide age range.

The present findings should be interpreted in light of potential self-selection bias; it is perhaps unsurprising that willingness to share mental health data was high amongst individuals who chose to complete a survey about the topic. That said, the data collected in our survey captured a small fraction of the information held in the health records about which we enquired. In addition, whilst the research sheds light on the impact of lifetime experience of mental illness on willingness to share mental health data, it does not identify those who were experiencing mental illness at the time of the survey. Further research should examine whether previous and current experience of mental illness have different impacts on data sharing. Similarly, future work would benefit from recruitment of more individuals with experience of less common mental health conditions in order to validate and extend the current findings. It would also be beneficial to assess whether public awareness campaigns aimed at reducing mental health stigma can eliminate the difference in willingness to share mental and physical health data in the UK.

## Conclusions

This study has shown for the first time that higher satisfaction with the NHS is associated with greater willingness to share mental health data. Furthermore, despite oft-cited concerns that mental health data are especially sensitive (4-8), it was found that people with experience of mental illness are actually more willing than people without mental illness to share their mental health data, once NHS satisfaction is taken into account. Much of the literature on preparing the ground for increased sharing of routinely-collected health data focuses on ways in which researchers can use public engagement to support greater buy-in from the general public (4). The present findings suggest that work to improve satisfaction with the NHS (29, 30) could also have a beneficial effect on willingness to share health data, especially amongst those with mental illness. In this way, both researchers and clinicians can play an important role in fostering greater public acceptability of research that uses routinely-collected mental health data. This in turn is expected to benefit researchers, clinicians and patients alike.

## Data Availability

The de-identified data are available at: doi.org/10.7488/ds/3146

## List of abbreviations

NHS: National Health Service
OCD: obsessive-compulsive disorder
OR: odds ratio

## Declarations

### Ethics approval and consent to participate

The research received ethical approval from the Department of Clinical and Health Psychology School of Health in Social Science Research Ethics Research Panel Committee at the University of Edinburgh (ref: STAFF132). All participants were presented with an online information sheet and provided informed consent using an online form at the beginning of the survey.

### Consent for publication

Not applicable

### Availability of data and materials

The datasets used and/or analysed during the current study are available at https://doi.org/10.7488/ds/3146.

### Authors’ contributions

EJK made contributed to the design of the work, conducted acquisition, analysis and interpretation of the data, and drafted the manuscript. SFW contributed to the conception and design of the work, the acquisition, analysis and interpretation of the data, and substantively revised the draft. AM contributed to the conception and design of the work, and the interpretation of the data. SML, CJC, SWYC and MHI contributed to the conception of the work. NDJ and JG contributed to the analysis and interpretation of the data. IB contributed to the acquisition of the data. All authors read and approved the final manuscript.

## Acknowledgements

We would like to thank the following individuals and organisations for their support with this work: Dr Mark Adams, Mahmud Al-Gailani, Dr Natalie Banner, Katie Breeze, Prof Rudolf Cardinal, Philippa Carr, Alison Churchill, Mark Coles, Judith Cramb, Clare Curtis, Dr Emma Davidson, Prof Ian Deary, Dr Josie Dickerson, the Edinburgh Pathfinder Stakeholder Advisory Group, Prof John Geddes, Dr Jane Haley, Prof Jeremy Hall, Emma Hawkins, Dr Sumeet Jain, Anders Jespersen, Prof Ann John, Prof Mark Johnson, Linda Jones, Prof Ian Jones, Catriona Keerie, Prof Simon Lovestone, Jonathan MacBride, Dr Donald MacIntyre, Prof John Macleod, Liz MacWhinney, Kate McAllister, Lanarkshire Links, Niamh McDade, Maria McMahon, Isabella Melking, Lynn Molloy, Denise Munro, National Survivor User Network, Jenny Nelder, Anne O’Donnell, Prof David Osborn, Prof Martyn Pickersgill, Prof David Porteous, Dr Carys Pugh, Emma Robinson, Dr Dan Robotham, Prof Ilina Singh, Prof Daniel Smith, Prof Robert Stewart, Suzy Syrett, Judith Syson, Adele Taylor, Leela Thomas, Liesbeth Tip, Shobna Vasishta, Hope Virgo, VOX Scotland, Prof James Walters, Katherine Woods.

